# COVID-19 policies in practice and their direct and indirect impacts in Northern California jails

**DOI:** 10.1101/2022.01.11.22269106

**Authors:** Yiran E. Liu, Christopher LeBoa, Marcela Rodriguez, Beruk Sherif, Chrisele Trinidad, Michael del Rosario, Sophie Allen, Christine Clifford, Jennifer Redding, Wei-ting Chen, Lisa G. Rosas, Carlos Morales, Alexander Chyorny, Jason R. Andrews

## Abstract

**Background:** Although the increased risk of COVID-19 in carceral facilities is well documented, little is known about the practical barriers to infection control and indirect impacts of pandemic policies in these settings. Evidence in jails is especially scarce.

**Methods:** Between July 8, 2020 and April 30, 2021 we performed SARS-CoV-2 serology testing and administered a questionnaire among residents and staff in four Northern California jails. We analyzed seroprevalence in conjunction with demographic factors and survey responses of self-perceived COVID-19 risk, recent illness, COVID-19 test results, and symptom reporting behaviors. We additionally assessed COVID-19 policies in practice and evaluated their impacts on court dates, mental health, and routine health care. We engaged stakeholder representatives, including incarcerated individuals and their advocates, to guide study design, conduct, and interpretation.

**Results:** We enrolled 788 incarcerated individuals and 380 staff across four county jails. Most seropositive individuals had not previously tested positive for COVID-19, despite many suspecting prior infection. Among incarcerated participants, we identified deficient access to face masks and prevalent symptom underreporting associated with fears of isolation and perceptions of medical neglect in jail. Incarcerated participants also reported substantial hindrances to court cases and reductions in routine health care due to COVID-19. Incarcerated individuals and staff both cited worsened mental health due to COVID-19, which for incarcerated individuals was largely attributable to further isolation from loved ones and other pandemic restrictions on recreation and programming.

**Conclusion:** Perceptions of inadequate protection from COVID-19 were pervasive among incarcerated individuals. Simultaneously, restrictive measures compounded poor mental health and fostered fears of isolation that undermined effective infection control. Custody officials should work to systematically improve provision of masks, understand and mitigate fears and mistrust, and take proactive steps to minimize the detrimental impacts of restrictive policies on residents’ mental health and well-being.

## INTRODUCTION

Mass incarceration has exacerbated the COVID-19 pandemic, with elevated infection and death rates among incarcerated individuals and staff [1-4] and mounting evidence of infection spillover from carceral facilities to outside communities [5-7]. Furthermore, incarceration disproportionately affects groups that already experience heightened vulnerability to COVID-19, including Black, Latinx, and Indigenous people, people experiencing unstable housing, and people with mental illnesses or substance use disorders [8-12].

In addition to exposing individuals to higher risk for COVID-19 morbidity and mortality, incarceration during the pandemic has also had indirect effects on people living and working in these facilities [13-18]. For instance, the CDC recommends that carceral institutions implement medical quarantine for exposed individuals and suspension of non-essential visitation and programming; these policies may magnify experiences of isolation for incarcerated individuals already removed from their communities [16, 19]. Moreover, the prolonged fear of exposure to COVID-19 may have had impacts on mental health for incarcerated individuals as well as for staff, especially those who were responsible for outbreak management and/or caring for incarcerated patients with COVID-19.

Despite the potentially profound direct and indirect impacts of the COVID-19 pandemic on the health and well-being of people living and working in carceral facilities, there has been limited primary, participatory research conducted in these settings, especially in jails. Compared to prisons, where all residents have been convicted and are serving sentences of years or more, jails have higher rates of population turnover, with 75% of residents in pre-trial detention and the remaining serving shorter sentences [10]. Jails also tend to have fewer resources and recreational opportunities for residents compared to prisons [20, 21]. Given these differences, the lack of data from county jail systems has been cited as an obstacle to evidence-based interventions in this unique setting [22, 23]. Moreover, research in carceral settings in general often forgoes consultation of community stakeholders and may consequently overlook relevant areas of investigation and local contextual factors important for study design and results interpretation.

The goal of this study was to understand the experience of the pandemic among individuals living and working in four county jails in Northern California using a stakeholder-engaged approach. These jails implemented various measures to mitigate COVID-19 spread, including testing and 14-day isolation of newly admitted individuals, 14-day quarantine of exposed individuals, provision of masks, and testing for residents and staff. In addition to examining these policies in practice and their indirect impacts on incarcerated individuals and jail staff, we performed SARS-CoV-2 serology testing and compared results with self-perceived risk and reported history of flu-like illness. Throughout the study, we engaged a community advisory board (CAB) consisting of incarcerated individuals, local and national advocates, public defenders, and custody health representatives to inform study design, execution, and analysis. Our findings underscore the importance of a holistic approach to infection control policies and may inform practices to improve the health and well-being of people living and working in jails during and beyond pandemic times.

## METHODS

### Overview

Between July 8, 2020 and April 30, 2021, we enrolled individuals living and working in four jails in San Mateo County and Santa Clara County, California to participate in SARS-CoV2 serology testing and complete a self-report questionnaire. This study was approved by the Stanford University and Valley Medical Center Institutional Review Boards (protocol #56169 and #20-022, respectively).

### Participant population and recruitment

In response to the pandemic, both San Mateo County and Santa Clara County implemented an emergency bail schedule and arrest reductions in order to de-densify their jails, resulting in jail populations of approximately 520 and 2000 incarcerated individuals, respectively. The staff population--including custody staff, healthcare workers, and program staff--remained relatively stable, with approximately 480 and 1050 staff members in San Mateo County and Santa Clara County, respectively. Incarcerated individuals were recruited through flyers and announcements in their housing units; in single-cell units, recruitment was done door-to-door. Research coordinators recruited from each housing unit in each jail at least once during the study period, with the exception of isolation units for COVID-19-positive individuals, units deemed by custody staff to be of high security risk, and units housing people with severe mental illnesses. Staff were recruited through flyers posted at work, emails sent by custody health officials, and radio announcements by custody leadership. Participation in any part of the study was voluntary with no penalty for refusal to participate. Due to timelines for administrative approvals, enrollment periods in each county were relatively distinct (**Fig S1)**.

### Serology test and seroprevalence analysis

Participants provided finger-prick blood samples for serology testing. Seropositivity was measured using the RightSign COVID-19 IgG/IgM Rapid Test Cassette manufactured by Hangzhou Biotest Biotech, granted emergency use authorization by the FDA on fingerstick blood samples [24]. All jails had an up-to-date CLIA certificate of waiver. In an internal validation of test performance using serum samples from a separate study of COVID-19-positive patients (true positives), the device had 81.5% and 92.1% sensitivity on patient samples from the day of and 28^th^ day following COVID-19 diagnosis, respectively [25]. Conversely, it had 100% specificity on 50 serum samples collected prior to 2019 (true negatives). Individuals who were vaccinated in custody prior to enrollment were excluded from seroprevalence analyses. Data on vaccination in jail were accessed as previously described [26]. Incarcerated participants received a hard copy of their serology results on the day of sample collection; staff received results over secure email within 1-2 days. All participants were provided with an informational flyer in English or Spanish on how to interpret their antibody test results and the option of consultation with study staff regarding their results.

### Questionnaire and other participant characteristics

Research coordinators administered the questionnaire using an electronic tablet. Separate questionnaires were developed for incarcerated participants and staff participants. Participants could choose, in English or Spanish, to read and respond to questions themselves or choose to respond orally to questions read aloud. Survey data were recorded in a HIPAA-secure REDCap database [27].

The questionnaire assessed demographic information, recent flu-like illness, perceptions and behaviors surrounding COVID-19, and indirect impacts of pandemic policies. For incarcerated participants whose demographic information (age, gender, race/ethnicity) or incarceration start date were missing, we accessed this information in their custody record (San Mateo County) or electronic health record [EHR] (Santa Clara County). Individuals were classified into the following racial/ethnic groups: Hispanic/Latinx, non-Hispanic white, non-Hispanic Black, non-Hispanic Asian, and non-Hispanic other or unknown race.

Questions on vaccination, medical trust, and sources of COVID-19 information were added in December 2020 for incarcerated participants and are analyzed in a separate paper [26].

### Community advisory board and stakeholder engagement

Two community advisory boards (CABs), one in each county, guided the overall design and implementation of this study. The goal of the CABs was to ensure that all parts of the study were relevant and sensitive to incarcerated individuals and community stakeholders. In each county, the CAB consisted of people who were currently incarcerated in the jails, representatives from custody health, a community organizer from a local advocacy organization, a national advocate for criminal justice reform, and in Santa Clara County, a public defender. Participation in the CAB was voluntary and non-binding. The Stanford study team met with each CAB periodically throughout the study via video conferencing, during which researchers provided an overview of the study aims and design and solicited feedback and suggestions about recruitment, ethical issues, survey questions, interpretation of results, and dissemination of findings. Meeting notes were circulated to CAB members following each meeting. In addition to CAB meetings, research staff hosted focus group discussions in various housing units within the jails prior to the enrollment start date as well as halfway through the study. These discussions were not limited to study participants and were intended to provide transparency into the research process, solicit feedback for the study, and address any questions or concerns regarding the study from stakeholders outside of the CABs. Key insights from CAB meetings and focus groups are presented throughout the Discussion.

### Statistical analysis

95% confidence intervals for seroprevalence were calculated using the Wilson method for binomial data [28]. Differences in seroprevalence and survey responses among groups were tested for significance using the chi-square test of independence. Correlation between seroprevalence and perceived likelihood of past infection was tested using the Spearman rank correlation test. All analyses were performed in R 4.0.5.

## RESULTS

### Study population

We enrolled 788 incarcerated individuals and 380 staff members across four jails in adjacent Northern California counties. This sample represented approximately 31% and 25% of the resident and staff population, respectively, across both counties combined. The incarcerated participant population was mostly male (89%), between the ages of 18 and 49 (85%), and Hispanic/Latinx or non-Hispanic Black (66%) (**Table 1**). The median length of incarceration at the time of enrollment was 80 days (IQR 15-285). Among staff participants, 38 % identified as health care workers. 28% of incarcerated participants and 29% of staff participants reported at least one medical condition considered a potential COVID-19 comorbidity.

**Table 1.**
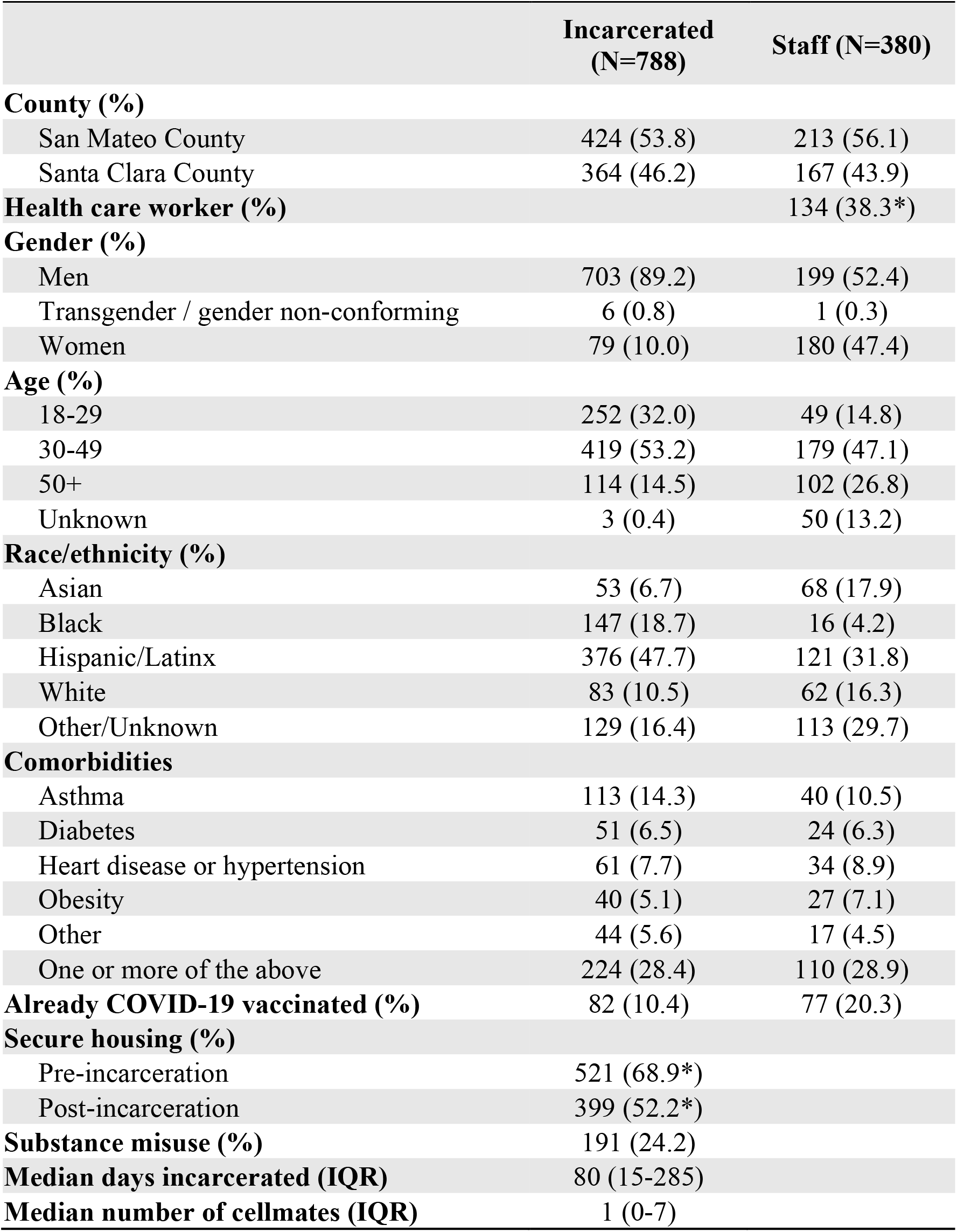
Characteristics of study participants. Percentages were calculated after excluding those with missing or “prefer not to answer” responses and may not sum to 100 due to rounding. Other comorbidities included cancer, immunosuppression, kidney or liver disease, chronic lung disease or COPD.

|  | **Incarcerated**<br>**(N=788)** | **Staff (N=380)** |
| --- | --- | --- |
| **County (%)** |  |  |
| San Mateo County | 424 (53.8) | 213 (56.1) |
| Santa Clara County | 364 (46.2) | 167 (43.9) |
| **Health care worker (%)** |  | 134 (38.3*) |
| **Gender (%)** |  |  |
| Men | 703 (89.2) | 199 (52.4) |
| Transgender / gender non-conforming | 6 (0.8) | 1 (0.3) |
| Women | 79 (10.0) | 180 (47.4) |
| **Age (%)** |  |  |
| 18-29 | 252 (32.0) | 49 (14.8) |
| 30-49 | 419 (53.2) | 179 (47.1) |
| 50+ | 114 (14.5) | 102 (26.8) |
| Unknown | 3 (0.4) | 50 (13.2) |
| **Race/ethnicity (%)** |  |  |
| Asian | 53 (6.7) | 68 (17.9) |
| Black | 147 (18.7) | 16 (4.2) |
| Hispanic/Latinx | 376 (47.7) | 121 (31.8) |
| White | 83 (10.5) | 62 (16.3) |
| Other/Unknown | 129 (16.4) | 113 (29.7) |
| **Comorbidities** |  |  |
| Asthma | 113 (14.3) | 40 (10.5) |
| Diabetes | 51 (6.5) | 24 (6.3) |
| Heart disease or hypertension | 61 (7.7) | 34 (8.9) |
| Obesity | 40 (5.1) | 27 (7.1) |
| Other | 44 (5.6) | 17 (4.5) |
| One or more of the above | 224 (28.4) | 110 (28.9) |
| **Already COVID-19 vaccinated (%)** | 82 (10.4) | 77 (20.3) |
| **Secure housing (%)** |  |  |
| Pre-incarceration | 521 (68.9*) |  |
| Post-incarceration | 399 (52.2*) |  |
| **Substance misuse (%)** | 191 (24.2) |  |
| **Median days incarcerated (IQR)** | 80 (15-285) |  |
| **Median number of cellmates (IQR)** | 1 (0-7) |  |

### Seroprevalence among incarcerated and staff participants by demographic characteristics and history or perceptions of prior infection

Among incarcerated participants, 13% (88/690; 95% CI, 11-16%) tested positive for antibodies to SARS-CoV2 (IgG and/or IgM) (**Table 2**). Among staff participants, 8% (24/292; 95% CI, 6-12%) tested positive. Seroprevalence was higher in Santa Clara County than San Mateo County among both incarcerated and staff participants (p<0.001). Seroprevalence also varied significantly by race/ethnicity of incarcerated participants (p=0.005), with the highest seroprevalence among Asian (24%) and Hispanic/Latinx (16%) individuals.

**Table 2.**
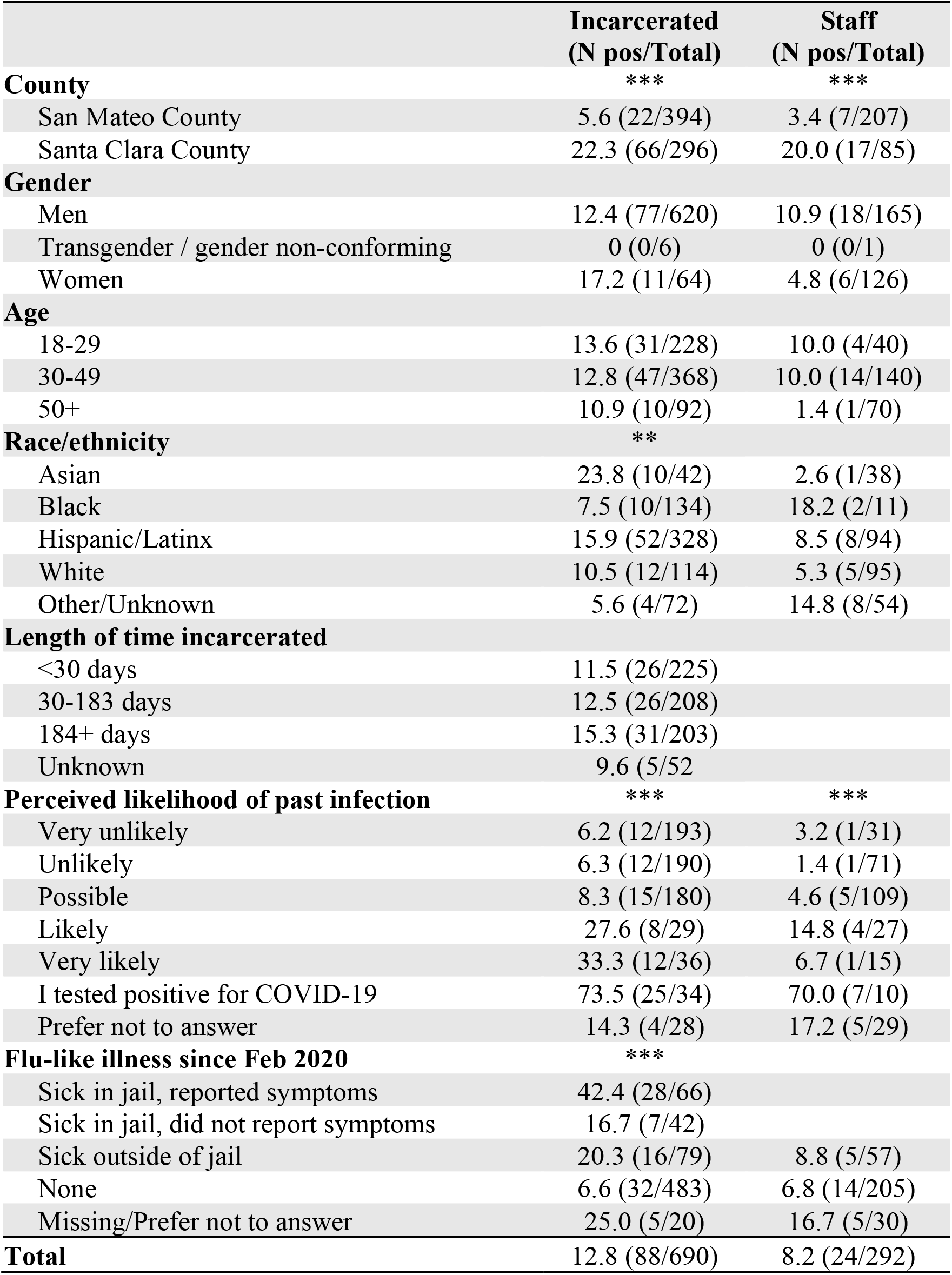
Seroprevalence by demographic characteristics and perceptions of past infection. Participants already vaccinated at the time of enrollment were excluded. Chi-square tests of independence were performed after excluding unknown, missing, or “prefer not to answer” responses. ** p<0.01; *** p<0.001.

Among incarcerated and staff participants who reported previously testing positive for COVID-19, 74% and 70% were positive for SARS-CoV2 antibodies, respectively (**Table 2**). Inversely, only 29% of seropositive participants reported previously testing positive for COVID-19. Among incarcerated participants who did not report a prior positive test, seroprevalence was significantly correlated with self-perceived likelihood of past infection (p=0.02). This trend was weaker among staff participants (p=0.13).

### Under-recognized and underreported illness among incarcerated participants and distrust toward jail staff

Approximately one-fourth of incarcerated participants (n=215, 27%) reported having a flu-like illness since February 2020, with 123 (16%) who said they had this illness in jail. Seroprevalence was significantly higher among incarcerated participants who said they had a flu-like illness, in or out of jail, compared to those who said they did not (p<0.001). Of those who were sick in jail, most (61%) reported their symptoms to jail staff, of which 62% and 52% said they were then tested for COVID-19 and/or put in isolation, respectively (**Fig 1A**). However, 22% indicated that no action was taken. For the 39% who said they did not report their symptoms to jail staff, the leading reason was not thinking it was not serious enough to report (47%), followed by not thinking anything would be done about it (28%), concern about being put in isolation (26%), and worry about how jail staff would treat them (21%) **(Fig 1B)**.

**Figure 1.**
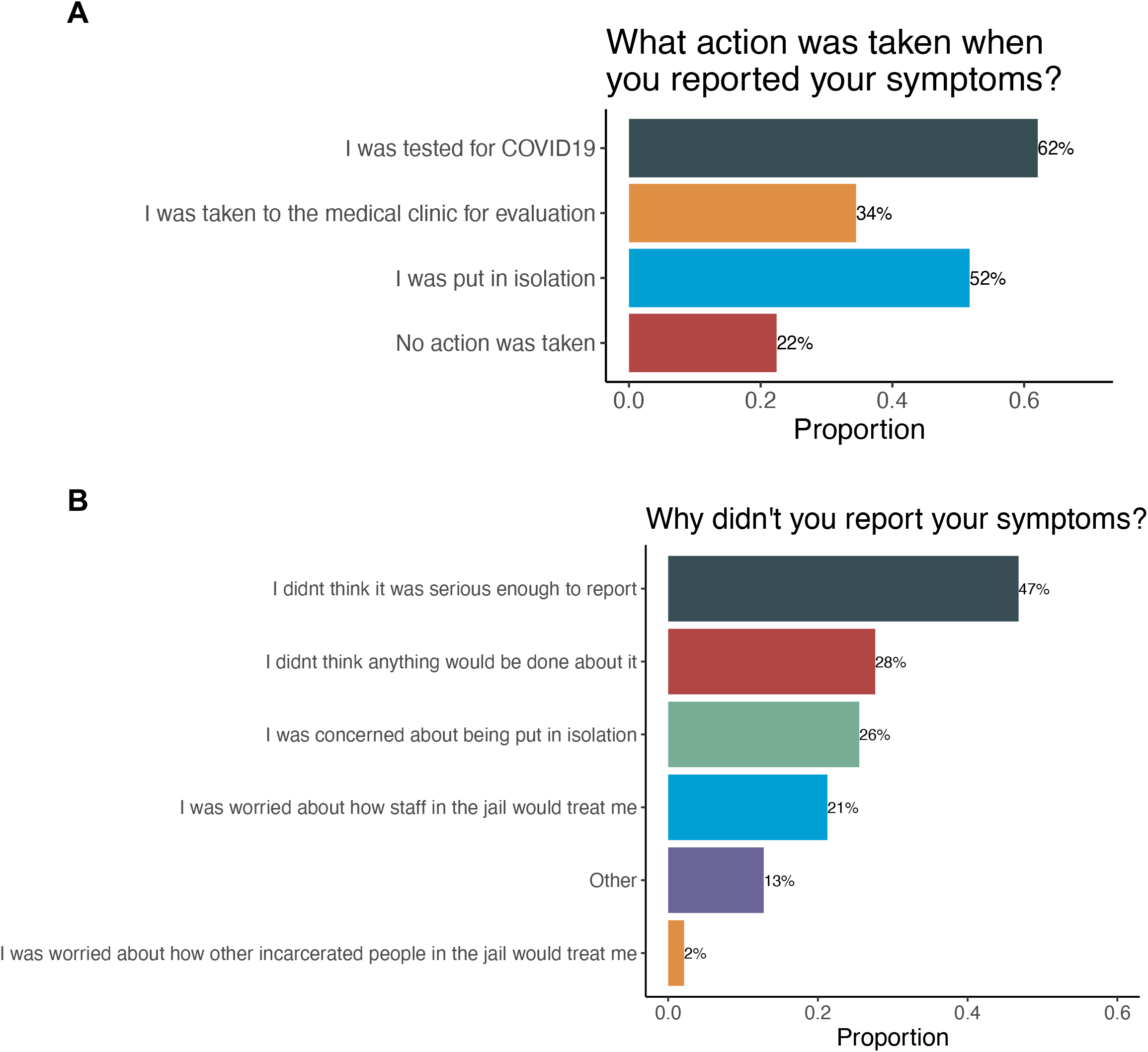
Actions taken following symptom reporting and reasons for underreporting among incarcerated participants. Among incarcerated participants with a flu-like illness in jail since February 2020, A) the 61% who reported their symptoms to jail staff were asked what actions were taken in response, and B) the remaining 39% were asked why they didn’t report their symptoms. Both questions were “select all that apply.”

Beyond underreporting of COVID-19-relevant symptoms, we identified general concerns around barriers to proper health care while in custody. When we asked incarcerated participants whether they believed that their health concerns were taken seriously by correctional officers or jail health staff, only 23% and 35% agreed with this statement regarding correctional officers and jail health staff, respectively **(Table S1)**. In comparison, 60% agreed that their health concerns were taken seriously by their doctor outside of jail. Moreover, 43% of incarcerated participants said they worried that they would be denied the medical treatment or services they needed while incarcerated, compared to 27% who had this same worry regarding their time outside of jail **(Table S1)**.

### Perceptions of protection from COVID-19 while in jail or at work

Among incarcerated participants, 54% reported feeling unable to protect themselves from COVID-19 while in jail, while only 5% of staff participants felt unable to protect themselves while at work **(Table S2)**. Incarcerated participants also reported frequent stress or fear around getting COVID-19 in jail, with 39% of incarcerated participants saying they experienced this fear often or all the time, compared to 20% of staff participants. However, 39% of staff participants did report frequent stress or fear around bringing COVID-19 from work to others in their household or community.

When incarcerated participants were asked to select three things that would protect them most from COVID-19, the leading response was release from jail (75%), followed by face masks/personal protective equipment (PPE) (56%) **(Fig S2)**. Among participants incarcerated for at least 30 days, 39% said they received a new mask less frequently than once a month, and another 33% reported having only received one mask since March 2020 (**Fig 2)**.

**Figure 2.**
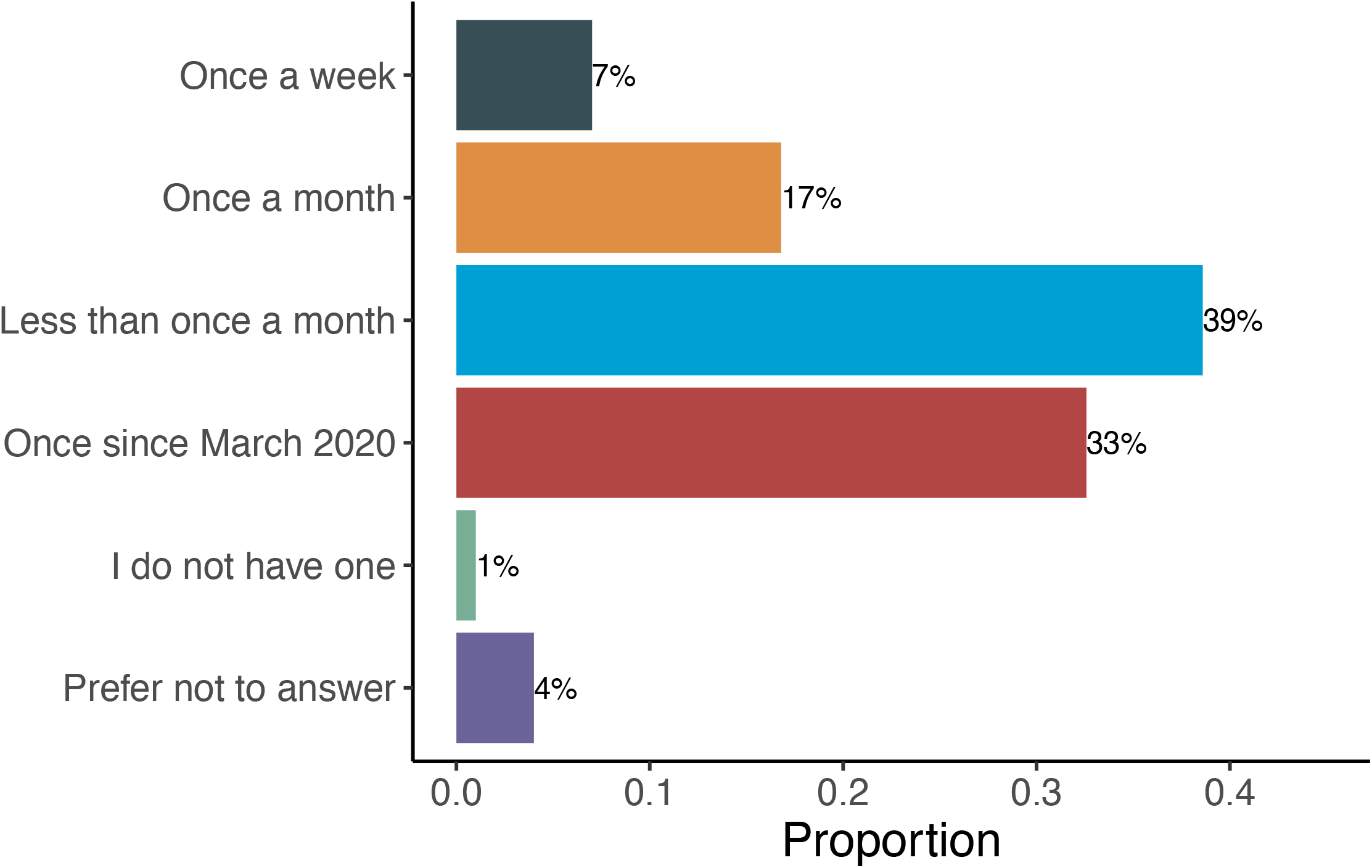
Reported frequency of receiving a new face mask among incarcerated participants. Participants incarcerated for less than 30 days were excluded. Percentages may not sum up to 100 due to rounding.

We next assessed whether participants believed that enough was being done to protect incarcerated individuals from COVID-19. 21% of incarcerated participants and 67% of staff participants agreed or strongly agreed with this statement **(Table S2)**. When asked whether they felt enough was being done to protect *staff* from COVID-19, 51% of staff participants agreed or strongly agreed.

### Impacts of COVID-19 on court dates, mental health, and routine health care

Among incarcerated participants, 61% indicated that their court dates were impacted by COVID-19. Of these, delays (76%), limits on attendance (56%), and cancellations (39%) were the most common impacts cited **(Fig S3A)**. Notably, of those whose court dates were delayed, 44% reported delays of over two months **(Fig S3B)**.

COVID-19 also had impacts on mental health, with 38.4% of incarcerated participants and 26% of staff participants citing worse mental health as a result of COVID-19. For incarcerated participants, the leading reasons were lack of connection to family and other loved ones (75%) and fear of getting COVID-19 (67%) **(Fig 3)**. Other common reasons included limits on programming (ie. classes, support groups) (56%), changes in recreation time (56%), unsanitary/unsafe conditions (56%), family or personal issues (55%), financial insecurity due to COVID-19 (46%), and lack of information about COVID-19 (39%). Among staff participants, reasons included fear of getting COVID-19 (64%), unsanitary/unsafe conditions at work (45%), family or personal issues (43%), lack of information about COVID-19 (26%), and frequency of COVID-19 routine testing (23%) **(Fig S4)**.

**Figure 3.**
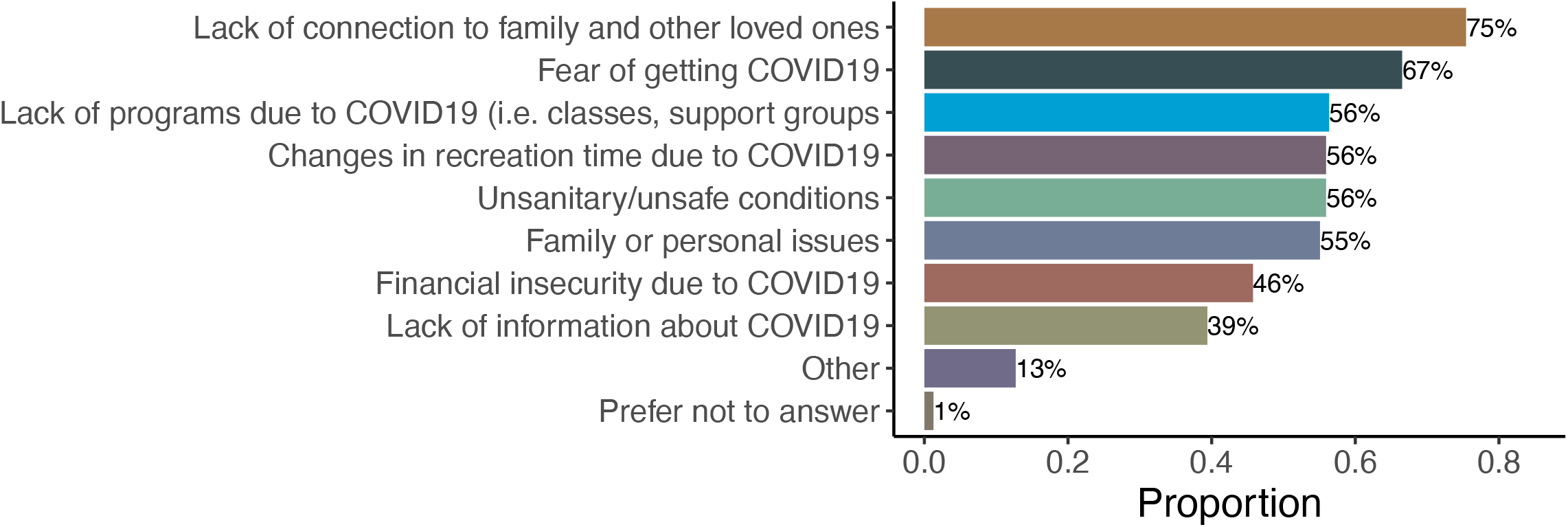
Reasons cited for worse mental health among incarcerated participants. eported factors affecting mental health (select all that apply) among the 43% of incarcerated participants who said their mental health has been worse or much worse due to COVID-19.

Finally, COVID-19 had indirect impacts on incarcerated individuals receiving routine mental or physical health care in jail. Of the 38% and 43% who reported previously receiving regular mental or physical health care in jail, respectively, approximately 40% said their health care had decreased or stopped due to COVID-19 **(Fig S5)**.

## DISCUSSION

Prisons, jails, and detention centers have been dangerous settings for COVID-19 transmission. There has been a paucity of research on perceptions and behaviors surrounding COVID-19 and the impacts of pandemic policies on the health and well-being of incarcerated individuals and staff, particularly in jails. In this study across four Northern California county jails, serology testing revealed a hidden burden of COVID-19 among incarcerated individuals, particularly among those who suspected prior infection but had not been tested. Our survey results identified several potential contributors to this, including symptom underreporting and perceptions of medical neglect. We also demonstrate the substantial indirect impacts of COVID-19 policies on incarcerated individuals through court delays, recreation and program restrictions, and further isolation from loved ones. Together, these findings shed light on practical barriers to infection control and surveillance in jails and underscore the importance of considering direct and indirect impacts of pandemic policies on health and well-being.

We found that most seropositive individuals had not previously tested positive for COVID-19, as has been shown in studies outside of carceral settings [29], and that these undetected infections were concentrated among incarcerated individuals who had a high self-perceived risk of prior infection. Accordingly, incarcerated members of our CAB noted that the jails’ policy of 14-day isolation for COVID-19 cases and known contacts--during which access to the phone, shower, recreation time, and programming could be further restricted--had stoked widespread perceptions among incarcerated individuals that a positive COVID-19 test would effectively lead to solitary confinement. This in turn, they explained, deterred symptom reporting, testing uptake, and other care-seeking behaviors, including attending routine health care appointments. These insights call for the clear, transparent communication of jail quarantine policies, as well as the critical need for procedures and practices in carceral settings that demonstrably distinguish the material conditions of medical isolation from those of solitary confinement [14, 16].

Another issue raised by stakeholder representatives was around residents’ access to face masks, which is one of few safeguards available given the difficulty of physical distancing in this setting. Current CDC guidance calls for “routine” washing and replacement of masks for incarcerated individuals but lacks explicit recommendations for implementation of such guidance [19]. Indeed, incarcerated focus group members expressed concern around being unable to safely wash and dry their mask without another to wear in the meantime. According to custody health officials, incarcerated individuals could ask custody staff or medical personnel to exchange their soiled or torn mask for a new one without limitation. However, our findings suggest that in practice, access to new masks was extremely limited and highly variable across individuals. Relatedly, and perhaps consequently, most incarcerated participants felt unable to protect themselves from COVID-19. Our findings emphasize the need for systematic implementation and oversight of mask availability and distribution in these settings.

Both counties surveyed here recognized the importance of jail de-densification to COVID-19 control and subsequently extended California’s emergency bail schedule for lesser charges, leading to population reductions of up to 45% [30, 31]. However, many who remained in custody experienced severe hindrances to their cases due to COVID-19, citing substantial delays and cancellations of court dates. These findings correspond to those of a recent investigation into the backlog of cases in California that has been exacerbated by COVID-19, which found that over a quarter of unsentenced jail residents in the two counties we studied had been in custody for more than a year awaiting arraignment, trial, or sentencing [32]. Beyond delays, our survey respondents also reported limits on court attendance, not only of witnesses but also of family and community members. As highlighted by our CAB, such attendance is essential for participatory defense, a community organizing model developed locally that engages family and community members in shaping a loved one’s case [33].

Our findings revealed additional indirect impacts of pandemic policies on the health and well-being of jail residents. All four jails, in line with other facilities around the country [34], suspended in-person visitation for over ten months, a policy that continues to be reinstated as new variants surge [35]. During this suspension, residents were provided with limited allowances of free phone and video calls; however, our findings suggest that these were inadequate substitutes to prevent the detrimental effects of disconnection from family and other loved ones on residents’ mental health. Other COVID-19 policies were also implicated in worsened mental health, including restrictions in recreation time and suspension of in-person programming. Furthermore, some participants reported reductions in routine mental and/or physical health care due to COVID-19, as has been shown in non-carceral settings [36, 37]. In the present study, this may have arisen from changes in custody health operations and treatment programs due to COVID-19, as have been reported in other carceral facilities [13], as well as from reduced care-seeking due to fears of isolation described above. Altogether, while COVID-19 has also had numerous indirect impacts on non-incarcerated individuals, our findings illustrate the compounding effects of pandemic policies in a population already experiencing high rates of mental illnesses and the physical and psychological stressors of incarceration.

This study also examined perceptions and attitudes surrounding COVID-19 among jail staff. While staff generally felt able to protect themselves from COVID-19 at work, some still reported unsanitary conditions, worsened mental health due to COVID-19, and frequent fears of getting infected at work and bringing it home. Of note, most staff agreed that enough was being done to protect incarcerated individuals from COVID-19, in stark contrast to the perceptions of incarcerated participants. One possible explanation for this divergence is that staff and residents may preferentially value different interventions. For instance, staff may favorably perceive contact-reduction measures such as quarantine and suspension of programs, while incarcerated individuals--having their mental health worsened and court cases delayed by such measures--may instead value interventions like provision of masks and cleaning supplies, which fulfill rather than deprive their agency. However, additional research is needed to further examine this discrepancy as well as other perceptions and impacts of COVID-19 among staff.

This study had several limitations. All survey data were subject to self-reporting biases. Our respondent population was likely a biased sample due to voluntary participation; moreover, the resident population consisted of those who were not released on emergency bail and therefore may not be representative of the resident population prior to COVID-19. Differences between counties may be confounded in part by the differential periods of enrollment in each county. Due to the decentralization of jail systems, our findings may have limited generalizability to residents and staff in other jail systems, as well as to those in other types of carceral facilities such as prisons and detention centers. Finally, while we were guided by the CAB in designing survey questions and interpreting results, our understanding remains limited by our methods and would be enhanced by future qualitative work.

## Conclusions

This study reveals barriers to the effectiveness of infection control policies in carceral settings, including the harm of isolation and the ensuing fears that deter symptom reporting and other care-seeking. Deficiencies in practice in mask distribution and the response to reported illness may contribute to perceptions of neglect among incarcerated individuals, which may in turn undermine testing and vaccination uptake [26]. Additionally, pandemic policies have resulted in further social isolation and heightened case-related stress that exacerbate poor mental health and the already distressing conditions of incarceration. Collectively, our findings demonstrate the urgent need for custody officials to investigate and address sources of fear and mistrust among incarcerated individuals and to prioritize continuation of family contact, programs, and services essential for health and well-being.

## Data Availability

Data from the present study are not currently available due to ethical and privacy concerns but may be made available in de-identified or aggregate form in the future, as allowed by the IRB.

## SUPPORTING INFORMATION

**Table S1. Concerns about barriers to health care in and out of custody among incarcerated participants**. Percentages may not sum up to 100 due to rounding.

**Table S2. Perceptions surrounding protection from COVID-19 among incarcerated and staff participants**. Percentages were calculated after excluding those with missing or “prefer not to answer” responses and may not sum up to 100 due to rounding.

**Figure S1. Participants enrolled each month, by county and population**. Incarc, incarcerated.

**Figure S2. Perceptions of the most effective protective measures from COVID-19 among incarcerated participants**. Participants were asked to select the three things they felt help protect them most from COVID-19.

**Figure S3. Impacts of COVID-19 on court dates of incarcerated participants**. A) Reported effects on court dates (select all that apply) among the 61% of incarcerated participants who indicated that their court dates had been impacted by COVID-19. B) Of those who said their court dates were delayed, the approximate length of the delays. Percentages may not sum up to 100 due to rounding.

**Figure S4. Reasons cited for worse mental health among staff participants**. Reported factors affecting mental health while working in a jail during COVID-19 (select all that apply), among the 26.2% of staff participants who said their mental health has been worse or much worse due to COVID-19.

**Figure S5. Reported changes in regular mental or physical health care due to COVID-19 among incarcerated participants**. Among incarcerated participants who reported previously receiving regular mental and/or physical health care in jail prior to COVID-19, percent who reported changes in A) mental health care and B) physical health care in jail due to COVID-19. Percentages may not sum up to 100 due to rounding.

## REFERENCES

1. Nowotny, K.M., K. Seide, and L. Brinkley-Rubinstein, Risk of COVID-19 infection among prison staff in the United States. BMC Public Health, 2021. 21(1): p. 1036.

2. Marquez, N., et al., COVID-19 Incidence and Mortality in Federal and State Prisons Compared With the US Population, April 5, 2020, to April 3, 2021. JAMA, 2021.

3. Saloner, B., et al., COVID-19 Cases and Deaths in Federal and State Prisons. JAMA, 2020. 324(6): p. 602–603.

4. Toblin, R.L. and L.M. Hagan, COVID-19 Case and Mortality Rates in the Federal Bureau of Prisons. American Journal of Preventive Medicine, 2021. 61(1): p. 120–123.

5. Reinhart, E. and D.L. Chen, Carceral-community epidemiology, structural racism, and COVID-19 disparities. Proceedings of the National Academy of Sciences, 2021. 118(21): p. e2026577118.

6. Reinhart, E. and D.L. Chen, Association of Jail Decarceration and Anticontagion Policies With COVID-19 Case Growth Rates in US Counties. JAMA Network Open, 2021. 4(9): p. e2123405–e2123405.

7. Sims, K.M., J. Foltz, and M.E. Skidmore, Prisons and COVID-19 Spread in the United States. American Journal of Public Health, 2021. 111(8): p. 1534–1541.

8. Couloute, L., Nowhere to Go: Homelessness among formerly incarcerated people. 2018: Prison Policy Initiative.

9. Jones, A.S. Wendy, Arrest, Release, Repeat: How police and jails are misused to respond to social problems. 2019: Prison Policy Initiative.

10. Sawyer, W.W. Peter, Mass Incarceration: The Whole Pie 2020. 2020: Prison Policy Initiative.

11. Schnittker, J., M. Massoglia, and C. Uggen, Out and Down: Incarceration and Psychiatric Disorders. Journal of Health and Social Behavior, 2012. 53(4): p. 448–464.

12. Wang, Q.Q., et al., COVID-19 risk and outcomes in patients with substance use disorders: analyses from electronic health records in the United States. Molecular Psychiatry, 2021. 26(1): p. 30–39.

13. Bandara, S., et al., Early Effects of COVID-19 on Programs Providing Medications for Opioid Use Disorder in Jails and Prisons. Journal of Addiction Medicine, 2020. 14(5).

14. Osgood, B., Weeks Without a Shower: Neglect Defines COVID-19 Containment in California Jails, in The Intercept. 2021.

15. Johnson, L., et al., Scoping review of mental health in prisons through the COVID-19 pandemic. BMJ Open, 2021. 11(5): p. e046547.

16. Cloud, D.H., et al., Medical Isolation and Solitary Confinement: Balancing Health and Humanity in US Jails and Prisons During COVID-19. Journal of General Internal Medicine, 2020. 35(9): p. 2738–2742.

17. Salonga, R., California courts temporarily waive bail for low-level offenses as part of COVID-19 response, in Mercury News. 2020.

18. Tavoschi, L., et al., Prevention and Control of COVID-19 in Italian Prisons: Stringent Measures and Unintended Consequences. Frontiers in Public Health, 2020. 8.

19. CDC Interim Guidance on Management of Coronavirus Disease 2019 (COVID-19) in Correctional and Detention Facilities. 2021.

20. Rosen, D. Justice Delayed in California Jails: Lengthy Pretrial Imprisonment Common. Prison Legal News, 2021.

21. Pohl, J.G., Ryan California Tried to Fix Its Prisons. Now County Jails Are More Deadly. ProPublica, 2019.

22. Komarla, A.C.I.-C.P., COVID-19 vaccination data in California jails: lessons from an imperfect model 2021.

23. Komarla, A., S.F.’s jails are leading California in COVID prevention. How are they doing it? No one knows, in San Francisco Chronicle. 2021.

24. Premier Biotech Inc FDA Grants EUA to Rapid COVID-19 Antibody Testing Product Distributed by Premier Biotech. 2020.

25. Jagannathan, P., et al., Peginterferon Lambda-1a for treatment of outpatients with uncomplicated COVID-19: a randomized placebo-controlled trial. Nature Communications, 2021. 12(1): p. 1967.

26. Liu, Y.E., et al., Factors associated with COVID-19 vaccine acceptance and hesitancy among residents of Northern California jails. medRxiv, 2021: p. 2021.11.22.21266559.

27. Stanford REDCap Electronic Data Capture Platform.

28. Wilson, E.B., Probable Inference, the Law of Succession, and Statistical Inference. Journal of the American Statistical Association, 1927. 22(158): p. 209–212.

29. Pollán, M., et al., Prevalence of SARS-CoV-2 in Spain (ENE-COVID): a nationwide, population-based seroepidemiological study. The Lancet, 2020. 396(10250): p. 535–544.

30. Salonga, R., Santa Clara County courts extend $0 bail order until 2022, in Mercury News. 2021.

31. Davis, L.I., Emergency Bail Schedule, S.M.C.S. Court, Editor. 2021.

32. Lewis, R. Waiting for Justice. CalMatters, 2021.

33. Silicon Valley De-Bug. About Participatory Defense. [cited 2021.

34. How Prisons in Each State Are Restricting Visits Due to Coronavirus. 2020: The Marshall Project.

35. San Mateo County jails implement safety protocols amid omicron surge. Climate Redwood City, 2022.

36. Lange, S.J., et al., Potential Indirect Effects of the COVID-19 Pandemic on Use of Emergency Departments for Acute Life-Threatening Conditions - United States, January-May 2020. MMWR Morb Mortal Wkly Rep, 2020. 69(25): p. 795–800.

37. Czeisler, M., et al., Delay or Avoidance of Medical Care Because of COVID-19-Related Concerns - United States, June 2020. MMWR Morb Mortal Wkly Rep, 2020. 69(36): p. 1250–1257.

