## Supplemental Tables for "COVID-19 policies in practice and their direct and indirect impacts in Northern California jails"

**Table S1. Concerns about barriers to health care in and out of custody among incarcerated participants.** Percentages may not sum up to 100 due to rounding.

|  | **I believe that ______ take(s) my health concerns seriously.** | | | **While _____, I worry that I will be denied the treatment or services I need.** | |
| --- | --- | --- | --- | --- | --- |
|  | **The correctional officers** | **The jail doctors and nurses** | **My doctor outside of jail** | **In jail/prison** | **Outside of jail/prison** |
| **Strongly Agree** | 7% | 10% | 20% | 18% | 8% |
| **Agree** | 16% | 25% | 40% | 25% | 19% |
| **Neutral** | 27% | 30% | 24% | 28% | 23% |
| **Disagree** | 20% | 16% | 6% | 16% | 30% |
| **Strongly Disagree** | 23% | 14% | 6% | 6% | 13% |
| **Prefer not to answer** | 6% | 6% | 5% | 6% | 7% |

**Table S2. Perceptions surrounding protection from COVID-19 among incarcerated and staff participants.** Percentages were calculated after excluding those with missing or “prefer not to answer” responses and may not sum up to 100 due to rounding.

|  | **Incarc %** | **Staff %** |
| --- | --- | --- |
| **How well do you feel you can protect yourself from COVID-19 while in jail / at work?** |  |  |
| Able or very well able | 16.8 | 51.5 |
| Somewhat able | 29.2 | 43.8 |
| Not really or not at all able | 54.0 | 4.8 |
| **How often have you experienced stress, fear, worry, and/or anxiety about getting COVID-19 in jail / at work?** |  |  |
| Often or all the time | 38.9 | 20.1 |
| Sometimes | 23.5 | 27.4 |
| Occasionally or never | 37.7 | 52.5 |
| **How often have you experienced stress, fear, worry, and/or anxiety about bringing COVID-19 infection from work to others in your household or in your community?** |  |  |
| Often or all the time |  | 39.3 |
| Sometimes |  | 25.8 |
| Occasionally or never |  | 34.9 |
| **Enough is being done to protect incarcerated individuals from COVID-19** |  |  |
| Strongly agree | 5.5 | 22.5 |
| Agree | 15.0 | 44.4 |
| Neutral | 21.3 | 21.9 |
| Disagree | 23.9 | 7.5 |
| Strongly disagree | 34.3 | 3.6 |
| **Enough is being done to protect jail staff from COVID-19** |  |  |
| Strongly agree |  | 14.9 |
| Agree |  | 36.3 |
| Neutral |  | 30.4 |
| Disagree |  | 13.4 |
| Strongly disagree |  | 5.1 |
