## Supplemental Figures for "COVID-19 policies in practice and their direct and indirect impacts in Northern California jails"

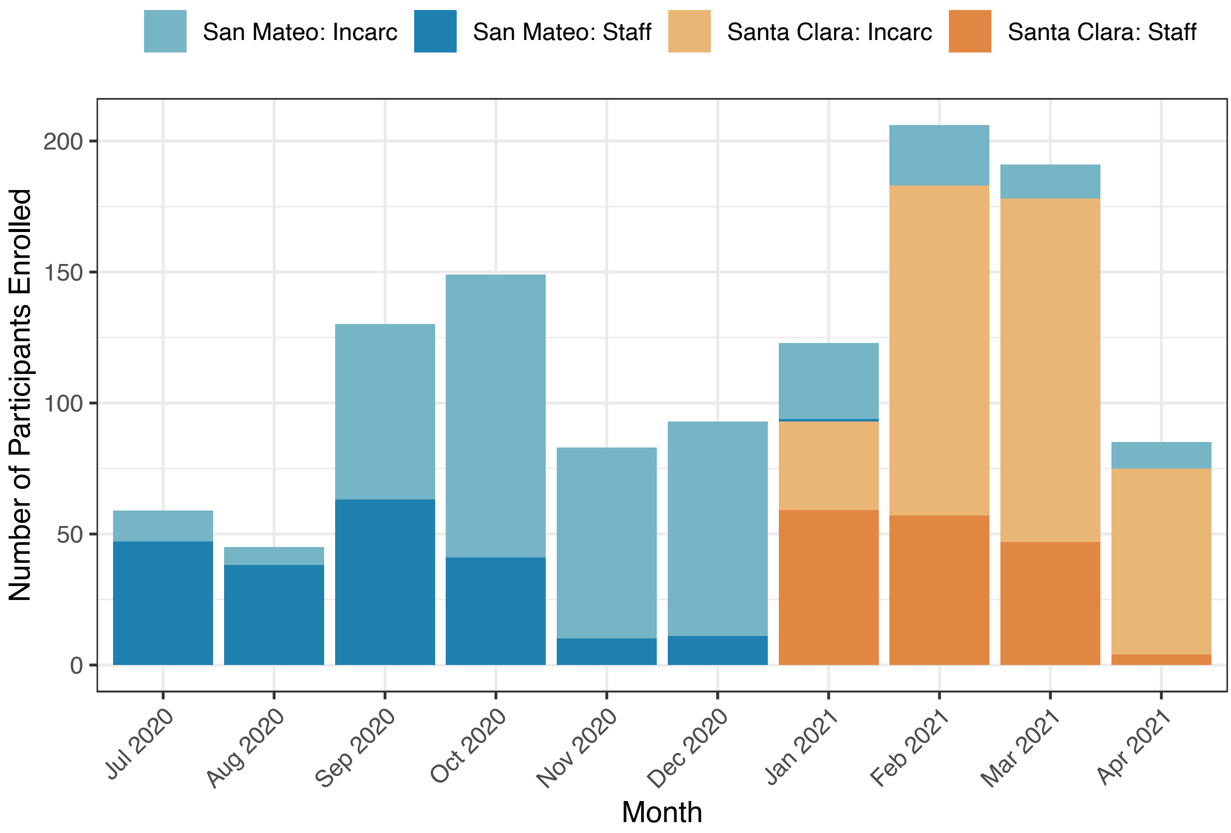


**Figure S1. Participants enrolled each month, by county and population.** Incarc, incarcerated.

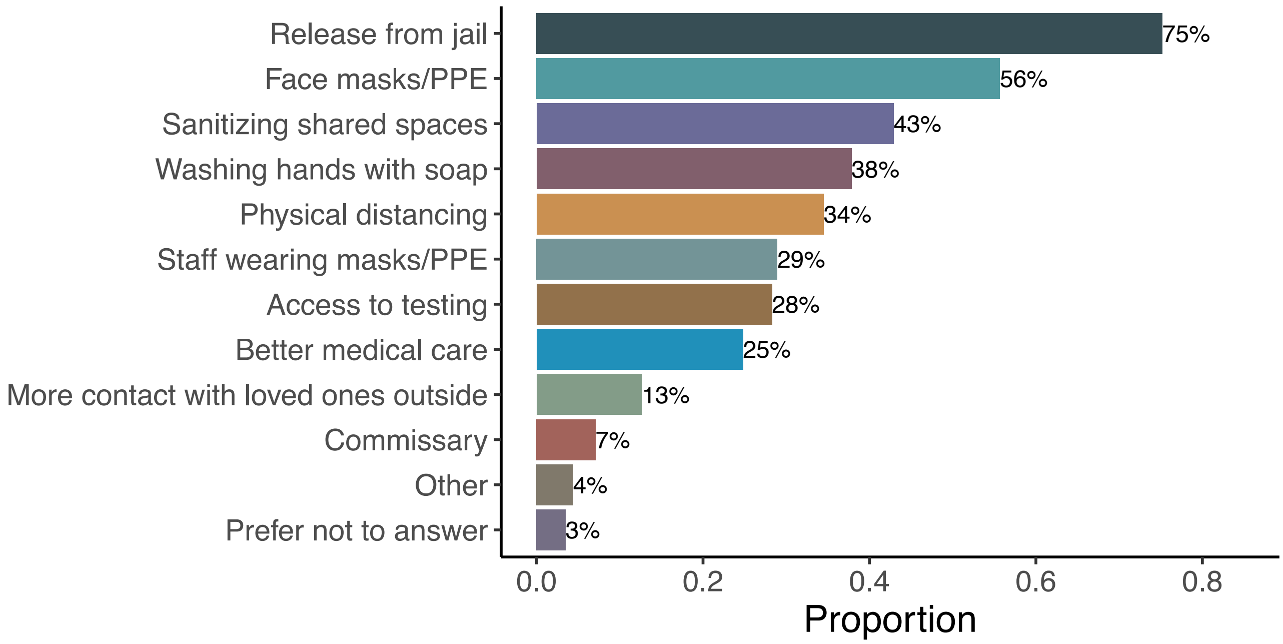


**Figure S2. Perceptions of the most effective protective measures from COVID-19 among incarcerated participants.** Participants were asked to select the three things they felt help protect them most from COVID-19.

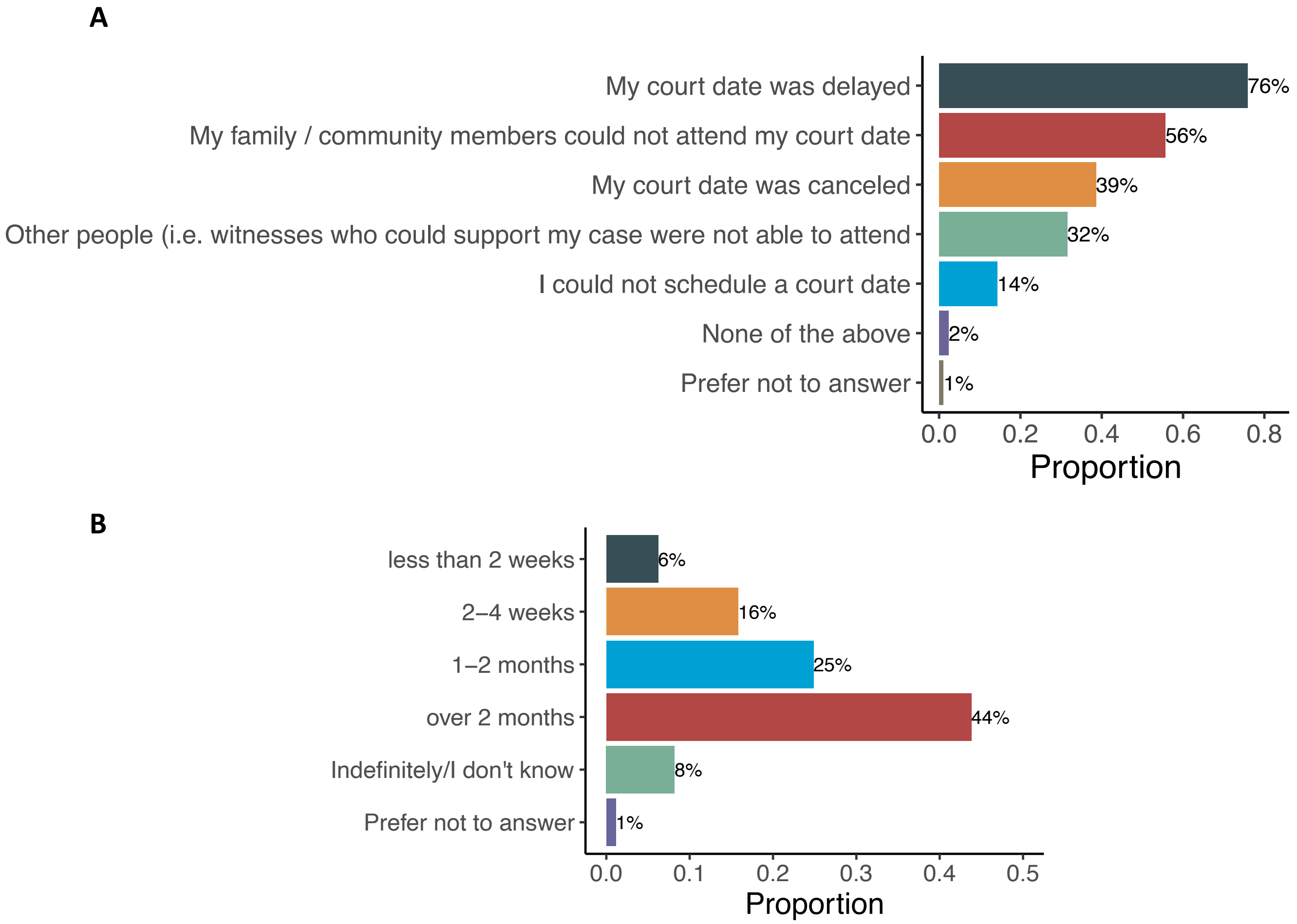


**Figure S3. Impacts of COVID-19 on court dates of incarcerated participants.** A) Reported effects on court dates (select all that apply) among the 61% of incarcerated participants who indicated that their court dates had been impacted by COVID-19. B) Of those who said their court dates were delayed, the approximate length of the delays. Percentages may not sum up to 100 due to rounding.

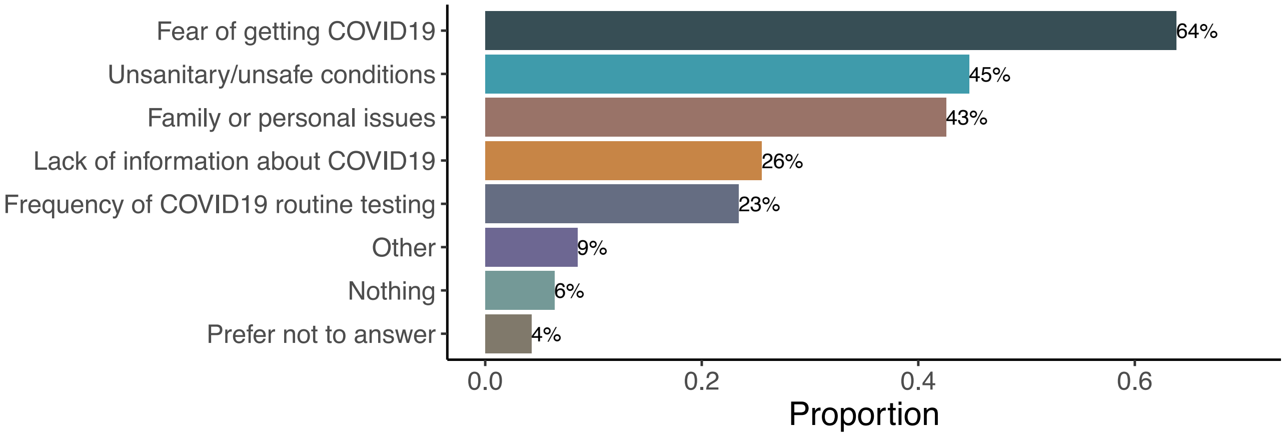


**Figure S4. Reasons cited for worse mental health among staff participants.** Reported factors affecting mental health while working in a jail during COVID-19 (select all that apply), among the 26.2% of staff participants who said their mental health has been worse or much worse due to COVID-19.

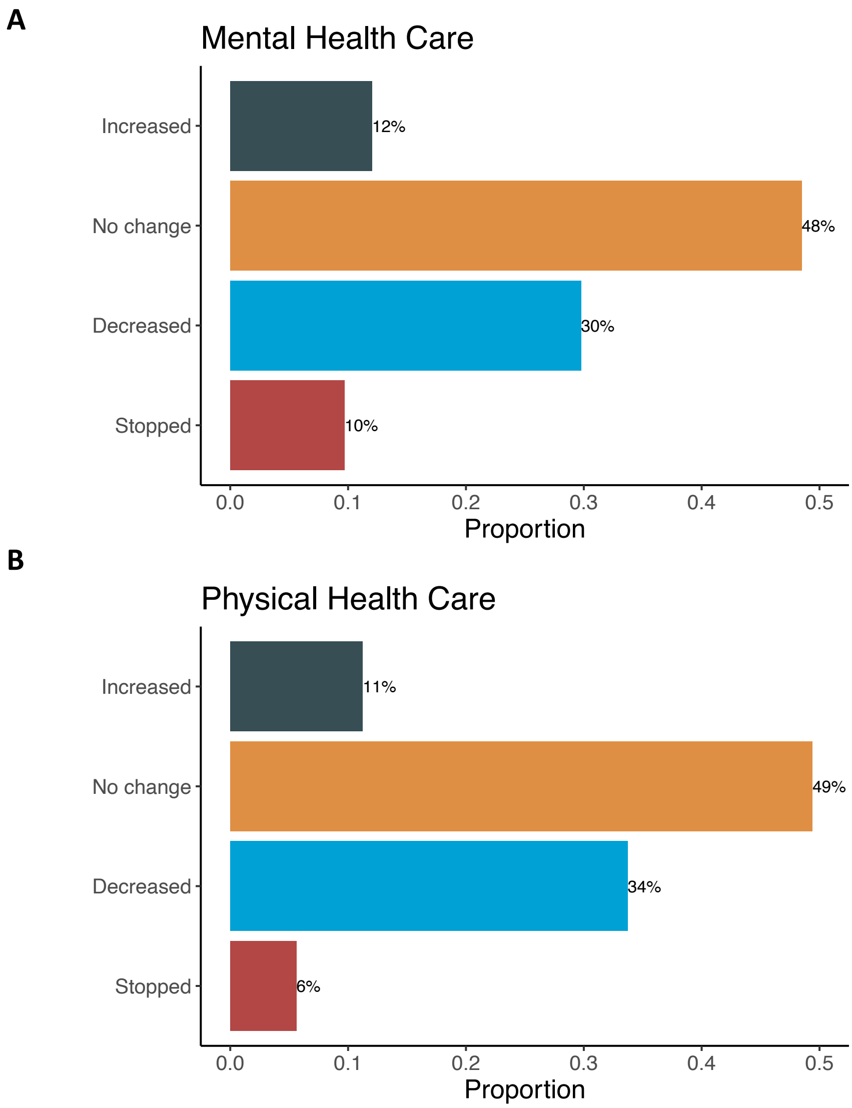


**Figure S5. Reported changes in regular mental or physical health care due to COVID-19 among incarcerated participants.** Among incarcerated participants who reported previously receiving regular mental and/or physical health care in jail prior to COVID-19, percent who reported changes in A) mental health care and B) physical health care in jail due to COVID-19. Percentages may not sum up to 100 due to rounding.
